# UM171-Expanded Cord Blood Transplantation in Adults with High- and Very High-Risk Acute Leukemia and Myelodysplastic Syndrome: Combined Results of Two Prospective Phase II Trials

**DOI:** 10.64898/2026.08.21.26360802

**Authors:** Sandra Cohen, Elisa Tomellini, Nadia Bambace, Imran Ahmad, Léa Bernard, Jean Roy, Jonathan Gutman, Jurjen Versluis, Pierre Caudrelier, Gabrielle Thauvette, Guy Sauvageau, Filippo Milano

## Abstract

**Purpose:** Adults with high- or very high-risk acute leukemia (AL) or myelodysplastic syndrome (MDS) face substantial relapse risk after allogeneic hematopoietic stem-cell transplantation. We evaluated single-unit cord blood (CB) transplantation after ex vivo expansion with UM171 in this population.

**Patients and Methods:** Two prospective, single-arm phase II trials at four centers enrolled 64 adults with high- or very high-risk AL or MDS; 60 received a UM171-expanded CB transplant and comprised the analysis population. CB units were preferentially selected at a 5/8 HLA match to maximize the graft versus leukemia effect. Patients received intermediate- or high-intensity conditioning with tacrolimus/mycophenolate mofetil graft-versus-host-disease (GVHD) prophylaxis. Endpoints included safety, feasibility, non-relapse mortality (NRM), relapse-free survival (RFS), overall survival (OS), GVHD, GVHD-free relapse-free survival (GRFS), chronic GVHD-free relapse free survival (CRFS).

**Results:** Thirty-two percent of patients had undergone previous transplantation, 17% of patients with AL were not in remission and 24% of those with AML/MDS had TP53 mutations. Of 62 patients who remained eligible for transplantation, 60 had a graft successfully manufactured and infused. Median times to neutrophil and platelet engraftment were 17 and 38 days, respectively. NRM was 5.1% at day 100 and 15.2% at 1 year. Two-year cumulative incidence of relapse was 22.3%. Two-year OS and RFS were 63.9% and 60.4%, respectively. Grade III–IV acute GVHD incidence was 20.3% at 1 year and moderate-to-severe chronic GVHD incidence was 6.8% at 2 years.

**Conclusion:** UM171-expanded CB transplantation was feasible and provided prompt engraftment, durable disease control, and infrequent clinically significant chronic GVHD in adults with high- and very high-risk AL/MDS. Comparative studies are warranted to define its role relative to contemporary donor platforms.

## Introduction

Relapse remains the leading cause of treatment failure after allogeneic hematopoietic stem-cell transplantation (allo-HSCT) for high- and very high-risk acute leukemia (AL) and myelodysplastic syndrome (MDS). Outcomes are particularly poor for patients with active disease at transplantation, TP53-mutated disease, or relapse after a previous allo-HSCT; in these settings, long-term survival rarely exceeds 20%-30% ^1–6^. Innovative strategies that enhance graft-versus-leukaemia (GvL) activity without increasing non-relapse mortality (NRM) and/or clinically significant graft-versus-host-disease (GVHD) are needed.

Cord blood (CB) is an attractive graft source for high-risk disease because of its distinctive immunological properties with several studies reporting lower relapse rates than with conventional donor sources in high-risk hematologic malignancies^7–10^. Moreover, CB permits greater HLA disparity, potentially augmenting GvL activity while expanding donor availability and maintaining a low incidence of chronic GVHD. However, conventional CB transplantation (CBT) is limited by low stem-cell dose, delayed haematopoietic recovery, a higher risk of graft failure, and increased NRM^11^. In this context, the restricted cell dose of most stored units often compels physicians to prioritise unit size over optimal HLA matching, potentially overlooking that increased HLA mismatch may itself contribute to higher NRM^12^. These limitations, combined with the widespread adoption of post-transplant cyclophosphamide (PTCy)-based haploidentical and mismatched unrelated donor transplantation have contributed to a marked decline in CBT over the past decade.

UM171 is a small molecule that promotes ex vivo self-renewal of human hematopoietic stem cells (HSC) and enables substantial expansion of CB-derived HSC while preserving long-term repopulating capacity^13^. In a phase I-II study, UM171-expanded CBT achieved rapid, sustained engraftment and enabled the use of smaller, better HLA-matched CB units, factors that likely contributed to the very low rate of NRM^14^.

Building on these results and evidence suggesting enhanced GvL activity with CBT, we conducted two prospective, multicentre phase II trials evaluating single-unit UM171-expanded CBT in adults with high- and very high-risk AL or MDS. CB units were preferentially selected at a 5/8 HLA match to enhance GvL activity. The trials were designed to confirm manufacturing feasibility, safety, and engraftment and to estimate relapse and survival outcomes in a population at high risk of treatment failure.

## Methods

### Study design

We conducted two parallel, single-arm, open-label phase II trials with minor differences in eligibility criteria, transplant procedures, and endpoint hierarchy. ECT-002 was a single center study conducted at Hôpital Maisonneuve-Rosemont (Montreal, Canada); ECT-004 was a multicenter study conducted at Fred Hutchinson Cancer Center (Seattle, WA), the University of Colorado School of Medicine, Anschutz Medical Campus (Aurora, CO) and Erasmus Medical Center Cancer Institute (Rotterdam, the Netherlands). ECT-002 was sponsored by Hôpital Maisonneuve-Rosemont, whereas ECT-004 was sponsored by ExCellThera Inc. Institutional review boards and regulatory authorities approved both protocols, and all patients provided written informed consent. The trials were registered at ClinicalTrials.gov (NCT03913026 and NCT04103879).

### Inclusion/Exclusion criteria

Adults age 18-70 years with high- or very high-risk AL or MDS were eligible. For AL, qualifying features included refractory disease, relapse after previous transplantation, adverse risk biology^15^, second complete remission (CR) or beyond, or measurable minimal residual disease before transplantation. MDS eligibility included relapse after previous transplantation, at least 10% blasts before conditioning, poor or very poor cytogenetics^16^, TP53 or any other high risk mutation, or lack of response to a hypomethylating agent. Patients were required to have a Karnofsky performance score of at least 70%, standard organ-function thresholds for transplantation and protocol-defined hematopoietic cell transplantation-comorbidity index (HCT-CI) limits based on age, disease status, and transplantation history. Complete protocol-specific criteria are provided in the Data Supplement.

### Graft selection and production

CB selection followed a prespecified algorithm requiring ≥4/8 HLA matching (HLA-A, -B, -C, -DRB1) with the patient, with a 5/8 match being the preferred match to maximize GvL activity. The selected unit required a minimum dose of 1.5×10^7^/kg total nuclear cells (TNC) and 0.5×10^5^/kg CD34^+^ cells. Units were shipped and processed at the Center of Excellence for Cellular Therapy at Hôpital Maisonneuve-Rosemont (Montreal, Canada).

After thawing, CD34-positive cells were selected and cultured with UM171 and growth factors for 7 days, then cryopreserved as drug product 1 (DP1). The CD34-negative fraction was cryopreserved as DP2. Manufacturing details are provided in the Data Supplement.

### Transplant Procedures

Three myeloablative conditioning regimens of intermediate or high intensity were permitted: (1) total body irradiation (TBI) 12-13.2 Gy, fludarabine (Flu) 75 mg/m^2^ and cyclophosphamide (Cy) 120 mg/kg); (2) TBI 4 Gy, Flu 150 mg/m^2^, Cy (50 mg/kg) and thiotepa (TT) 10 mg/kg^17^; (3) intravenous busulfan 6.4-9.6 mg/kg, Flu 150 mg/m^2^ and TT 10 mg/kg. GVHD prophylaxis consisted of tacrolimus (target 10–15 µg/L from D-3 through D+50, followed by taper and discontinuation by D+100) and mycophenolate mofetil 45 mg/kg/day (D-3 to D+35)^14^. DP1 and DP2 were infused on day 0. To mitigate any potential risk, the infused CD34^+^ cell dose was capped at 5.75 ×10^6^/kg and later amended to 7.5 ×10^6^/kg. Granulocyte colony-stimulating factor was administered until neutrophils ≥ 1.0 × 10⁹/L for 2 days. Infection prophylaxis followed institutional guidelines. Post transplant monitoring is detailed in the supplement.

### Endpoints

The two trials evaluated the same core efficacy and safety endpoints. RFS and OS at 1 and 2 years were primary efficacy endpoints in both trials. In ECT-002, the primary safety endpoint was NRM at day 100 and 1 year. In ECT-004, the primary safety endpoint included adverse events, graft failure, and feasibility of manufacturing and infusing a UM171-expanded CB graft, whereas NRM was a secondary endpoint. Combined secondary endpoints included engraftment, acute and chronic GVHD, grade ≥3 infections, pre-engraftment or engraftment syndrome requiring treatment, GVHD-free relapse-free survival (GRFS), chronic GVHD-free relapse-free survival (CRFS), and hospitalization events. Immune reconstitution was an exploratory endpoint of ECT-002. Post hoc analyses included relapse/progression incidence and exploratory evaluation of associations between age and other clinical or graft characteristics and NRM or relapse. Protocol-specific end-point definitions are provided in the Data Supplement.

### Statistical analysis

Baseline characteristics were summarized as medians with ranges or interquartile ranges (IQRs) and counts with percentages. NRM, defined as death without prior relapse or progression, and relapse/progression were estimated using cumulative incidence functions, treating each as a competing event for the other. OS was measured from transplantation to death from any cause. RFS was measured from transplantation to relapse/progression or death among patients in CR before transplantation or at the first post-transplant assessment; patients who never achieved CR were excluded. OS and RFS were estimated using the Kaplan-Meier method.

Neutrophil and platelet engraftment and acute and chronic GVHD were estimated using cumulative incidence functions, with death and relapse/progression as competing risks. Neutrophil engraftment was defined as the first of 3 consecutive days with neutrophil count ≥0.5 × 10⁹/L, and platelet engraftment as the first day of a sustained platelet count ≥20 × 10⁹/L without transfusion in the preceding 7 days. GRFS events included relapse/progression, death, grade III-IV acute GVHD, or chronic GVHD requiring systemic immunosuppression; CRFS included the same events except grade III-IV acute GVHD. GRFS and CRFS were estimated using the Kaplan-Meier method.

Exploratory analyses evaluated OS, RFS, NRM and relapse/progression according to age, assessed using both the cohort median (43 years) and prespecified categories of <40, 40–59, and ≥60 years.

The association between grade II-IV acute GVHD and relapse was evaluated using a cause-specific Cox model with acute GVHD as a time-dependent covariate. Analyses were performed using R version 4.0.3 or later. Outcomes are reported through 2 years and all patients completed the planned 2-year follow-up.

## Results

### Patients and CB Characteristics

Between May 2019 and December 2023, a total of 64 patients with high- or very high-risk AL or MDS were enrolled on both phase II prospective studies. Of these, 60 patients (94%) proceeded to HSCT (30 in ECT-002 and 30 in ECT-004) and constituted the study population. Four patients did not receive the study graft because of disease progression before transplantation (n = 2), shipping failure (n = 1), or failure to meet product release criteria (n = 1). As per protocol, these patients were removed from the study.

Patient and transplant characteristics were comparable between the two trials (**Table 1**). Median age was 43 years (range 19–66); 33 (55%) patients had AML, 19 (32%) had ALL, and 8 (13%) had MDS. Nineteen patients (32%) had previously undergone transplantation (18 allogeneic, 1 autologous). Among the 52 patients with AL, 9 (17%) were not in CR or incomplete CR (CRi)^15^ at transplant, and 20 (38%) were in CR2 or CR3. Amongst the 41 patients with AML or MDS, 10 (24%) had TP53 mutations and 2 (5%) had EVI1 rearrangements. Thirteen patients (22%) had an HCT-CI score of at least 3. Thirty-one patients (52%) were CMV-seropositive, of whom 23 received letermovir prophylaxis.

**Table 1.** Patient and transplant characteristics.

|  | Overall<br>(n=60) | ECT-002<br>(n=30) | ECT-004<br>(n=30) |
| --- | --- | --- | --- |
| Age in years, median (range) | 43 (19-66) | 48 (19-66) | 42 (21-64) |
| Sex, n(%) |  |  |  |
| Male | 37 (61.7%) | 22 (73.3%) | 15 (50.0%) |
| Female | 23 (38.3%) | 8 (26.7%) | 15 (50.0%) |
| Diagnosis n (%) |  |  |  |
| Acute Myeloid Leukemia (AML) | 33 (55.0%) | 16 (53.3%) | 17 (56.7%) |
| Acute Lymphoid Leukemia (ALL) | 19 (31.7%) | 8 (26.7%) | 11 (36.7%) |
| Myelodysplastic Syndrome (MDS) | 8 (13.3%) | 6 (20.0%) | 2 ( 6.7%) |
| Disease status at screening for AL, n (%) |  |  |  |
| Number of AL patients | 52 | 24 | 28 |
| CR1 | 23 (44.2%) | 10 (41.7%) | 13 (46.4%) |
| ≥CR2 | 20 (38.5%) | 8 (33.3%) | 12 (42.9%) |
| Not CR/CRi | 9 (17.3%) | 6 (25.0%) | 3 (10.7%) |
| Previous transplant, n(%) | 19 (31.7%) | 10 (33.3%) | 9 (30.0%) |
| allogeneic | 18 (30.0%) | 10 (33.3%) | 8 (26.7%) |
| autologous | 1 ( 1.7%) | 0 ( 0.0%) | 1 (3.3%) |
| Other high-risk biology features |  |  |  |
| TP53 mutation | 10 (16.7%) | 7 (23.3%) | 3 (10.0%) |
| EVI1 rearrangement | 2 (3.3%) | 2 (6.7%) | 0 (0.0%) |
| MRD positivity before transplant | 9 (15.0%) | 3 (10.0%) | 6 (20.0%) |
| Karnofsky performance score, n(%) |  |  |  |
| 90-100 | 35 (58.3%) | 11 (36.7%) | 24 (80.0%) |
| ≤80 | 25 (41.7%) | 19 (63.3%) | 6 (20.0%) |
| HCT-CI, median (range) | 1 (0-5) | 2 (0-4) | 1 (0-5) |
| Patients with HCT-CI score ≥3, n(%) | 13 (21.7%) | 9 (30.0%) | 4 (13.3%) |
| Conditioning regimen, n(%) |  |  |  |
| TBI 13.2 or 12 Gy/Flu/Cy | 14 (23.3%) | 6 (20.0%) | 8 (26.7%) |
| TBI 4 Gy/Flu/Cy/Thiotepa* | 43 (71.7%) | 21 (70.0%) | 22 (73.3%) |
| Bu/Flu/Thiotepa | 3 (5.0%) | 3 (10.0%) | 0 (0.0%) |
| CMV seropositive patients, n(%) | 31 (51.7%) | 7 (23.3%) | 24 (80.0%) |

\*One patient received TBI 2 Gy/Flu/Cy/Thiotepa due to pre-existing liver toxicity
Abbreviations: AL, acute leukemia; CR, complete remission; MRD, measurable residual disease; HCT-CI, Hematopoietic Cell transplantation- Comorbidity Index; TBI, total body irradiation; Gy, gray; Flu, fludarabine; Cy, cyclophosphamide; Bu, busulfan.

CB unit characteristics are summarised in **Table 2**. As specified by the study design, which favored selection of a 5/8 HLA-matched CB unit, 45 patients (75%) received a 5/8 matched CB, whereas 15 (25%) received a 6/8 or 7/8 matched CB. Before expansion, almost half (43%) of the CB units did not meet the American Society for Transplantation and Cellular Therapy (ASTCT) recommended minimum cell dose thresholds for single-unit CBT^18^. Median CD34-positive expansion after UM171 exposure was 46-fold, yielding a median viable CD34-positive cell dose of 39.2 × 10^5^/kg (IQR, 23.8-57.5 × 10^5^/kg). The protocol-defined CD34-positive dose cap was applied in 16 patients (27%). The median viable CD3-positive cell dose in DP2 was 2.2 × 10^6^/kg (IQR, 1.6-3.3 × 10^6^/kg).

**Table 2.** Expanded grafts characteristics.

|  | **Overall**<br>**(n=60)** | **ECT-002**<br>**(n=30)** | **ECT-004**<br>**(n=30)** |
| --- | --- | --- | --- |
| **CB unit HLA match, n (%)** |  |  |  |
| 5/8 | 45 (75.0%) | 22 (73.3%) | 23 (76.7%) |
| 6/8 | 11 (18.3%) | 7 (23.3%) | 4 (13.3%) |
| 7/8 | 4 (6.7%) | 1 (3.3%) | 3 (10.0%) |
| **Pre-expansion CB Unit Cell Doses (provided by CB bank), median (IQR)** |  |  |  |
| CD34 x 10<sup>5</sup>/kg | 2.13 (1.43-2.73) | 2.53 (1.56-3.15) | 1.76 (1.28-2.22) |
| TNC x 10<sup>7</sup>/kg | 2.67 (2.14-3.68) | 2.67 (2.17-3.64) | 2.69 (2.11-3.69) |
| **Expanded Fraction (DP1) Formulation at Infusion, n (%)** |  |  |  |
| Fresh | 2 (3.3%) | 2 (6.7%) | 0 (0.0%) |
| Cryopreserved | 58 (96.7%) | 28 (93.3%) | 30 (100%) |
| **Expanded Fraction (DP1) Graft Cell Doses, median (IQR)** |  |  |  |
| vCD34 x 10<sup>5</sup>/kg | 39.15 (23.78-57.50) | 48.60 (33.60-57.50) | 30.65 (19.18-47.83) |
| vTNC x 10<sup>7</sup>/kg | 0.50 (0.28-0.65) | 0.59 (0.42-0.66) | 0.41 (0.26-0.54) |
| **Unmanipulated Fraction (DP2) Graft Cell Doses, median (IQR)** |  |  |  |
| vCD3 x 10<sup>6</sup>/kg | 2.21 (1.59-3.31) | 2.68 (1.79-3.44) | 2.08 (1.51-2.78) |
| vTNC x 10<sup>7</sup>/kg | 1.54 (1.13-2.00) | 1.89 (1.41-2.36) | 1.26 (1.06-1.70) |

DP1 and DP2 doses listed are those after manipulation and prior to final cryopreservation.
Abbreviations: CBU, cord blood unit; IQR, interquartile range; TNC, total nucleated cells; vTNC, viable TNC; vCD34, viable CD34+ cells; DP, drug product. DP1 and DP2 cell doses were measured after manipulation and, for cryopreserved products, before final cryopreservation.

### Engraftment

Neutrophil engraftment occurred in 55 of 60 patients (92%), as two patients had primary graft failure, one died of sepsis on D+18, and two experienced early disease progression. The median time to neutrophil engraftment was 17 days (IQR, 14–21; **Figure 1A**), and the median time to achieve a neutrophil count of 100/µL was 10 days (IQR, 9–13). One graft failure occurred in the setting of high-titer donor-specific anti-HLA antibodies and the second followed low post-thaw CD34+ recovery (before expansion), poor expansion, and human-herpesvirus 6 (HHV-6) viremia despite complete donor chimerism. Platelet engraftment occurred in 53 of 60 patients (88%) at a median of 38 days (IQR, 32–44; **Figure 1B**). Seven patients did not achieve platelet engraftment because of early NRM, disease progression, or graft failure. Long-term engraftment was sustained, with no case of secondary graft failure or late cytopenias. Full donor chimerism was rapidly established, with a median of 100% donor myeloid (CD33^+^) and T-cell (CD3^+^) chimerism at D+14, which remained stable throughout follow-up (**Figure 1C**).

**Figure 1.**
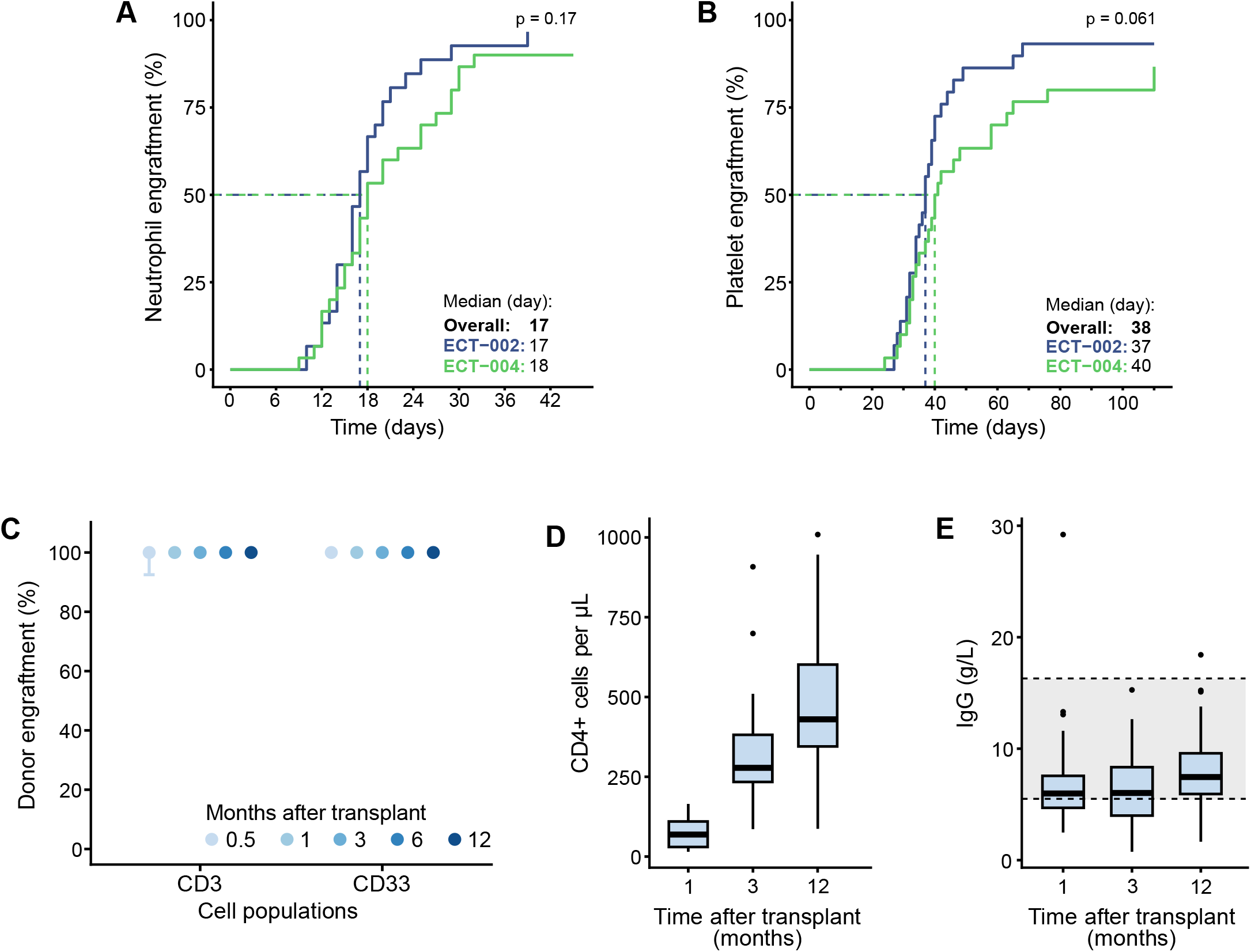
Engraftment kinetics and immune reconstitution following UM171-expanded cord blood transplantation. (A-B) Cumulative incidences of neutrophil (A) and platelet (B) engraftment in the ECT-002 and ECT-004 cohorts. (C) Donor chimerism in CD3^+^ and CD33^+^ cell populations at 0.5, 1, 3, 6, and 12 months post-transplant. (D) CD4^+^ T-cell counts at 1, 3 and 12 months post-transplant. (E) Immunoglobulin G (IgG) levels at 1, 3 and 12 months post-transplant; the shaded area indicates the normal range 5.5–16.3 g/L. Box plots show medians and interquartile ranges; whiskers extend to the most extreme value within 1.5 times the interquartile range.

### Clinical Outcomes

OS was 76.3% (95% CI, 63.3–85.2%) at 1 year and 63.9% (95% CI, 50.2–74.8%) at 2 years post-transplant (**Figure 2A**). RFS was 75.0% (95% CI, 61.5–84.4%) at 1 year and 60.4% (95% CI, 46.3–71.9) at 2 years (**Figure 2B**). Outcomes were similar between trials. Amongst patients younger than the median age of 43 years, outcomes were particularly favorable, with 2-year OS and RFS of 83% and 78%, respectively (**Figures 3A-B**).

**Figure 2.**
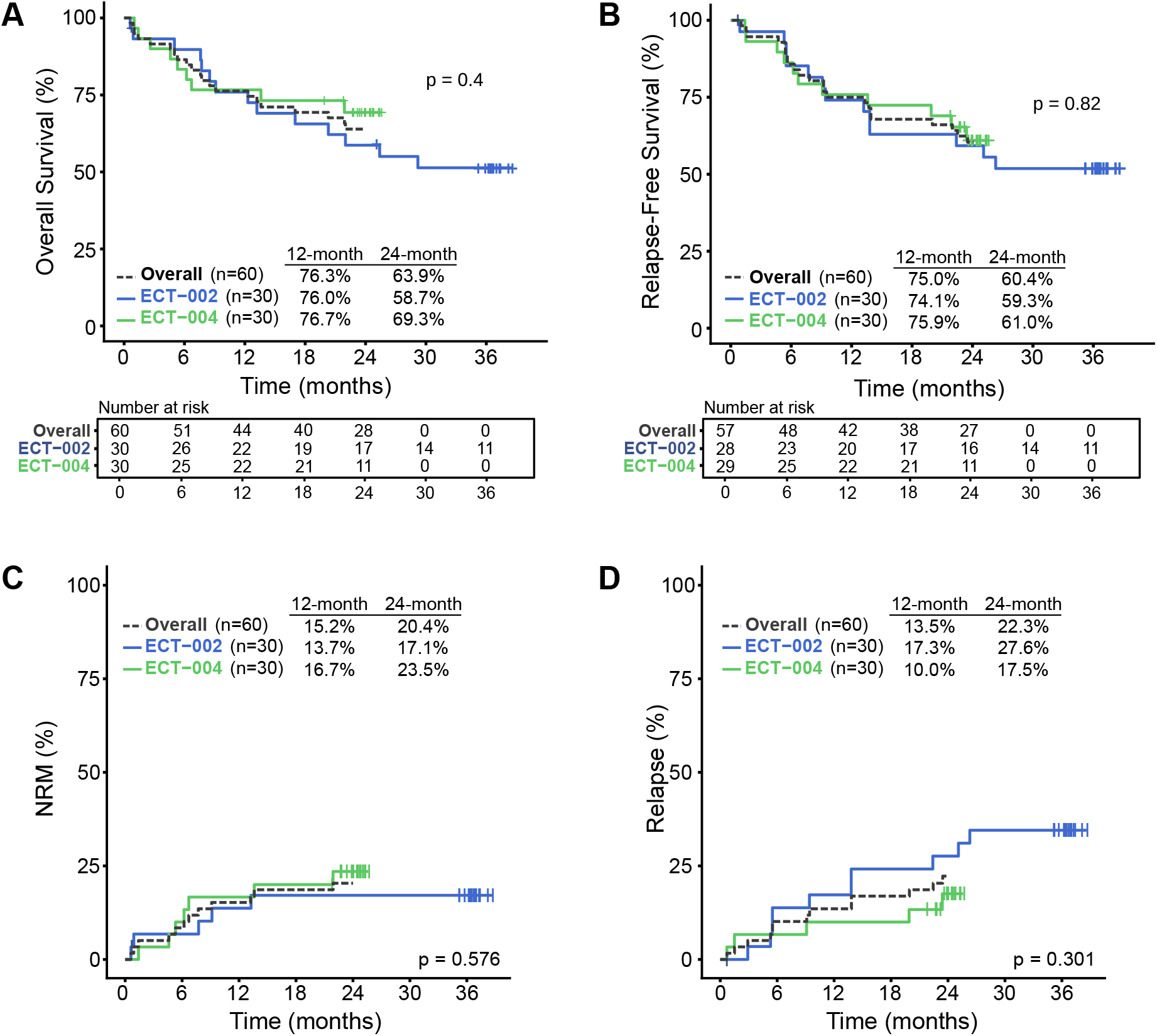
Survival outcomes, non-relapse mortality and relapse following UM171-expanded cord blood transplantation. (A-B) Kaplan-Meier estimates of overall survival (OS; A) and relapse-free survival (RFS; B). (C-D) Cumulative incidences of non-relapse mortality (NRM; C) and relapse (D). Tick marks indicate censored observations and numbers at risk are shown below each panel. P values were calculated using the log-rank test for OS and RFS and Gray’s test for NRM and relapse. ECT-002 patients were followed for 3 years and ECT-004 patients for 2 years.

**Figure 3.**
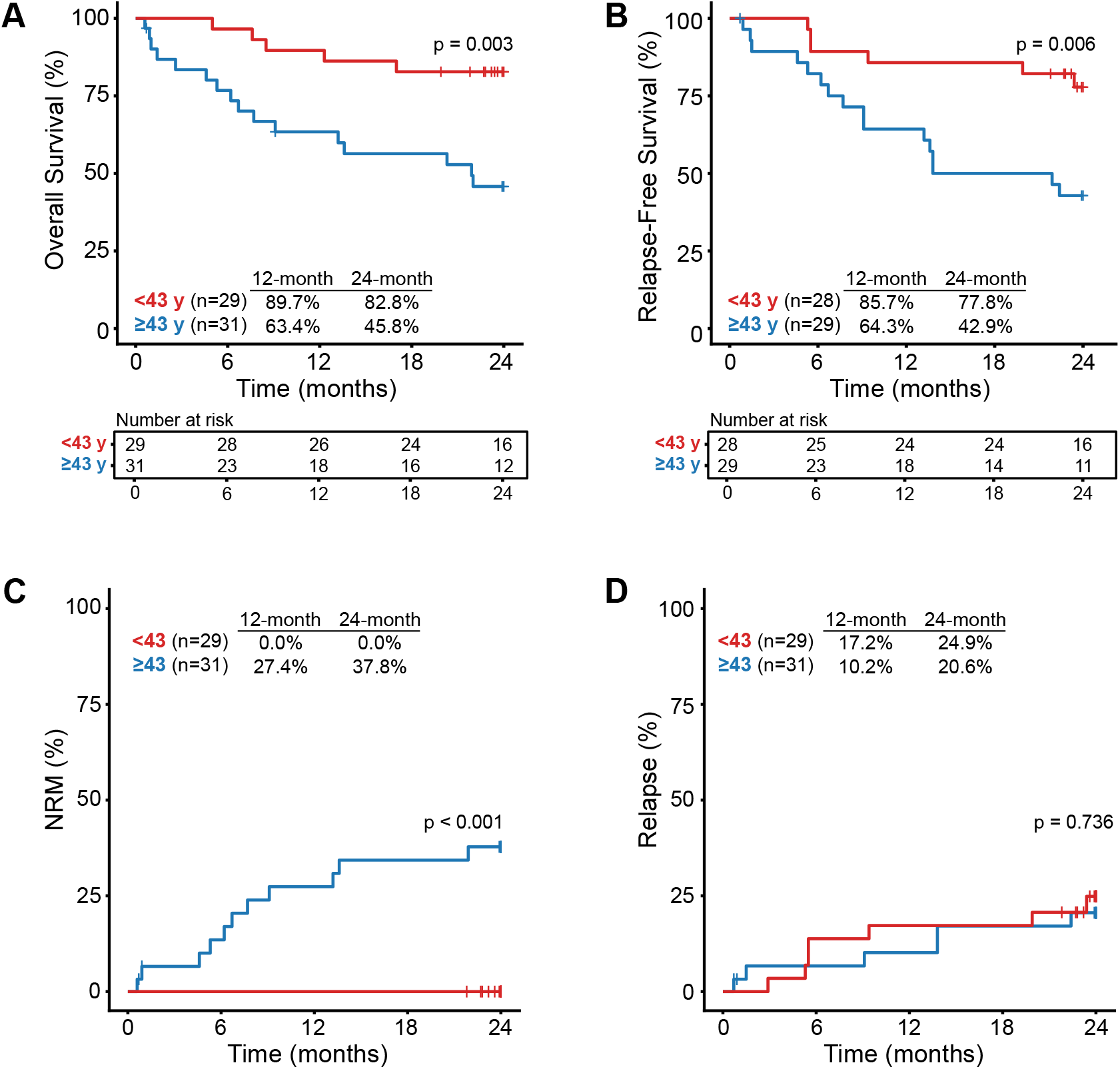
Outcomes following UM171-expanded cord blood transplantation stratified by age (<43 v ≥43 years). (A-B) Kaplan-Meier estimates of overall survival (OS; A) and relapse-free survival (RFS; B). (C-D) Cumulative incidences of nonrelapse mortality (NRM; C) and relapse (D). Tick marks indicate censored observations, and numbers at risk are shown below each panel. P values were calculated using the log-rank test for OS and RFS and Gray’s test for NRM and relapse.

The cumulative incidence for NRM was 5.1% (95% CI, 0.0–10.7%) at D+100 and 15.2% (95% CI, 6.0–24.5%) at 12 months (**Figure 2C**). Twelve NRM deaths occurred: 5 were attributed to infection, 4 to non-infectious pulmonary complications, 1 to multiorgan dysfunction, 1 to refractory acute GVHD, and 1 to graft failure (**Supp. Table 1**). In exploratory analyses, age was the only factor associated with NRM: 1-year NRM was 0% in patients aged <43 years versus 27% in those aged ≥43 years (P < .001; **Figure 3C**), although it was highest among patients aged 40–59 years when age was evaluated in three categories (**Supp. Figure 1**).

The cumulative incidence of relapse was 13.5% (95% CI, 4.7–22.3%) at 1 year and 22.3% (95% CI, 11.5–33.1%) at 2 years (**Figure 2D**) with no significant difference by age (**Figure 3D and Supp. Figure 1B**). In an exploratory time-dependent analysis, grade II-IV acute GVHD was associated with a lower hazard of relapse (HR, 0.32; 95% CI, 0.12-0.85; P = 0.02).

The cumulative incidence of grade II–IV acute GVHD at 1 year was 71.2% (95% CI 59-83%), with grade III–IV in 20.3% of patients (95% CI 10-31%), (**Figure 4A**). However, there was only one patient with steroid-refractory acute GVHD. At 2 years, the cumulative incidence of chronic GVHD was low at 12.2% (95% CI 4-21%), and that of moderate-to-severe chronic GVHD was very low at 6.8% (95% CI 0-13%) (**Figure 4B**). At 2 years, GRFS was 41.3% (95% CI, 30–56%), and CRFS 51.5% (95% CI, 38–64%) (**Supp. Figures 2A-B).**

**Figure 4.**
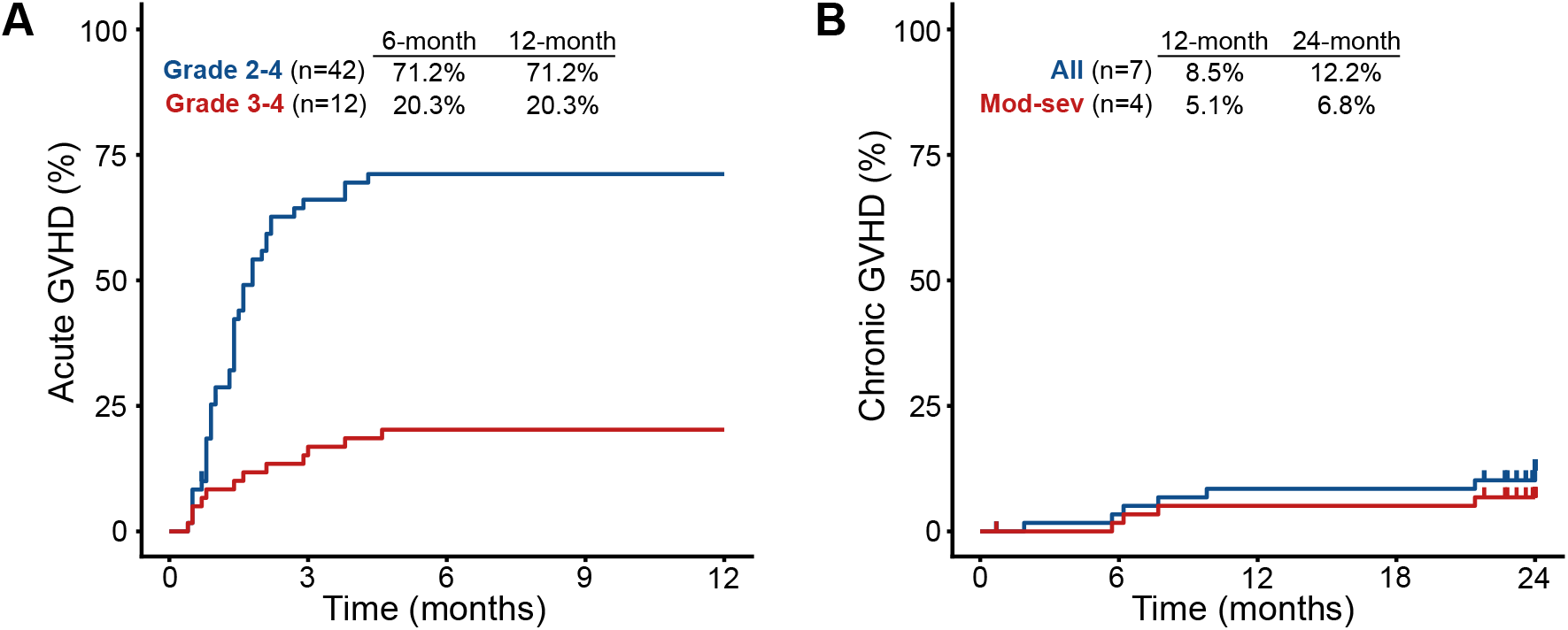
Graft-versus-host disease following UM171-expanded cord blood transplantation. (A) Cumulative incidence of acute graft-versus-host disease (GVHD), stratified by grade 2–4 and grade 3–4. (B) Cumulative incidence of chronic GVHD, including overall and moderate-to-severe (mod-sev) disease. Estimates at 6 and 12 months for acute GVHD and at 12 and 24 months for chronic GVHD are shown.

Outcomes in selected high-risk subgroups are shown in **Supp. Figure 3**. Amongst patients with TP53-mutated disease (n=10) and those undergoing a second allo-HSCT (n=18), RFS at 2 years was 50.0% (95% CI, 15.2–77.5%) and 44.4% (95% CI, 21.6–65.1%), respectively.

### Early Post-Transplant Outcomes and Immune Reconstitution

The median duration of hospitalization after transplant was 28 days (IQR, 19–36). Immune reconstitution was evaluated in ECT-002. Median CD4^+^ T-cell counts were 69, 278 and 430 cells/µL at 1, 3, and 12 months, respectively (**Figure 1D**). Median IgG levels remained within the normal range throughout follow-up; **Figure 1E**).

Grade 3 or higher adverse events are summarized in **Supplementary Table 2**. Among 31 CMV-seropositive patients, eight developed CMV reactivation, with no CMV disease. Seven patients received treatment for EBV viremia, with no post-transplant lymphoproliferative disorder. Four patients were treated for HHV-6 viremia.

## Discussion

In two prospective phase II trials enrolling adults with high- and very high-risk AL and MDS, a population in whom long-term survival rarely exceeds 20–30% after conventional allo-HSCT^1–6^, single-unit UM171-expanded CBT achieved a two-year OS of 64% and RFS of 60%, with a low relapse incidence of 22%. The cohort included patients with TP53-mutated disease, active disease at transplant, and previously failed allogeneic transplantation, all associated with particularly poor outcomes. Although cross-study comparisons require caution, these results compare favorably with published outcomes using contemporary donor platforms^1,2,4,5^.

Prompt and durable engraftment was achieved despite nearly half of CB units falling below minimum cell dose thresholds^18^. The 17-day median neutrophil engraftment time was consistent with our prior phase I–II experience^14^ and is comparable to conventional donor platforms^19,20^. The absence of secondary graft failure or late cytopenias supports the preservation of long-term hematopoietic stem-cell repopulating capacity after UM171 expansion. By overcoming the cell-dose constraint of conventional CBT, expansion may permit greater emphasis on HLA matching, particularly in standard-risk disease, where minimizing NRM may outweigh the potential GvL benefit of greater HLA disparity. In our phase I/II trial, expansion enabled selection of better-matched CB units in most patients and was associated with an NRM of 5%^14^. Robust immune reconstitution, including rapid CD4^+^ T-cell recovery, further supports graft quality; this is clinically relevant because delayed CD4^+^ recovery has been reported after PTCy-based transplantation^21,22^ whereas early CD4^+^ reconstitution is associated with improved survival^23 24^.

The 1-year NRM of 15% is noteworthy given the clinical complexity of this cohort and compares favorably with historical rates of 20%–30% in similarly high-risk populations undergoing conventional transplantation, with even higher rates in patients with TP53-mutated disease and those undergoing a second allo-HSCT^25–28^. No NRM events occurred among patients younger than 43 years translating into an excellent 2-year RFS of 78%. The observation that NRM was highest among patients aged 40–59 years suggests that chronological age alone did not explain this outcome. Several noninfectious pulmonary deaths occurred among older patients and second allo-HSCT recipients, raising the possibility of conditioning-related toxicity, particularly from TBI. Less toxic approaches, including lower dose TBI, treosulfan-based conditioning, and non-TBI regimens such as thiotepa-busulfan-fludarabine, should be tested in these higher risk groups^29^.

The 2-year relapse incidence of 22% represents an important efficacy signal, particularly given the adverse disease features of the study population and published relapse rates of 35-60% in comparable populations^1,2,4,30^. An enhanced GvL effect with CB transplantation has been suggested in several large retrospective studies, particularly in high-risk hematologic malignancies^7–10^. The biological basis for this effect remains incompletely understood but may reflect the distinctive immune properties of CB, including the capacity of naïve CD8^+^ T cells to rapidly acquire tumor-directed cytotoxicity without sustained alloreactivity^31,32^. The low relapse incidence in this study suggests that this activity is preserved after UM171 expansion.

Grade II-IV acute GVHD occurred in 71% of patients. Although this incidence is higher than that typically reported with PTCy- or ATG-based prophylaxis, it is consistent with rates reported after conventional ATG-free CBT and therefore does not suggest excess acute GVHD attributable to the UM171 expansion^33–36^. Only one patient developed steroid-refractory acute GVHD. The lower relapse risk in patients developing acute GVHD is consistent with the established association between GVHD and GvL activity.^37^ Most importantly, the 2-year incidence of moderate-to-severe chronic GVHD was only 6.8%, comparing favorably with rates reported with PTCy or ATG prophylaxis with conventional donors^35,36^. This chronic GVHD profile is important, as chronic GVHD remains a major cause of late morbidity and impaired quality of life after allo-HSCT. Thus, UM171-expanded CBT was characterized by frequent but generally treatment-responsive acute GVHD and infrequent clinically significant chronic GVHD.

In exploratory analyses, 2-year RFS was 50% among 10 patients with TP53-mutated AML/MDS and 44% among 18 second allo-HSCT recipients, compared with historical estimates of 15%–30% and 20%–35%, respectively^1,2,4,25–27,38,39^. Although limited by small samples and the absence of a concurrent control group, these findings support prospective evaluation in these high-risk populations.

Study strengths include its prospective multicenter design, complete 2-year follow-up, and consistent results across two independently conducted trials. Limitations include the modest sample size, single-arm design, heterogeneity in diagnosis, disease status, and conditioning, minor protocol differences, and the post hoc nature of some analyses. The study was not powered for subgroup comparisons.

In summary, UM171-expanded CBT enabled prompt, durable engraftment from small CB units while preserving GvL potential. Rapid engraftment, low NRM, encouraging disease control, and infrequent moderate-to-severe chronic GVHD represent a clinically distinctive profile warranting randomized comparison with contemporary alternative-donor platforms in adults with high-risk AL/MDS.

## Data Availability

All data produced in the present study are available upon reasonable request to the authors

## Acknowledgement

The Canadian study was funded by the Stem Cell Network (FY17/CT1), and Canadian Cancer Society Research Institute (CCSRI 703199). We would like to acknowledge Catherine Paquin, research nurse, for all her work pertaining to this study and the members of the Center of Excellence for Cell Therapy at Maisonneuve-Rosemont Hospital and C3i for manufacturing of the UM171 expanded cord blood grafts. We thank Hema-Quebec for their contribution with cord blood selection. Finally, we would like to thank the data safety monitoring board for their time.

## Author Contributions

Conception and design: SC conceived and designed ECT-002, with contributions from JR, IA, NB, and LB. FM contributed to the conception and design of ECT-004.

Collection and assembly of data: SC oversaw data collection for ECT-002; NB and IA reviewed and audited the source data. FM oversaw data collection for ECT-004, with JG and JV overseeing data collection at their respective sites. ET and GT merged the datasets from both trials.

Data analysis and interpretation: ET and GT performed the statistical analyses. SC, ET and GS performed the primary data analysis. SC led the interpretation of the results, with contributions from all authors.

Manuscript writing: SC wrote the initial draft of the manuscript. ET subsequently edited the manuscript, and SC incorporated comments and revisions from all authors into the final manuscript.

Final approval of manuscript: All authors. Accountable for all aspects of the work: All authors.

## Data Supplement

### Inclusion and Exclusion Criteria

#### ECT.002

##### Inclusion criteria

1. Presence of a high-risk acute leukemia/myelodysplasia as defined by one of the following:

a. Acute Myelogenous Leukemia

i. Primary induction failure.
ii. Chemorefractory relapse.
iii. Relapse after autologous or allogeneic transplant.
iv. High risk AML in CR1: any adverse genetic abnormality as defined by ELN excluding FLT3 mutation, secondary or therapy related AML excluding good risk genetic abnormalities, or any poor risk feature known to be associated with a PFS or DFS ≤40% at 2 years after a conventional transplant.
v. CR2 excluding good risk genetic abnormalities.
vi. ≥CR3
b. Acute Lymphoblastic Leukemia

i. Primary induction failure.
ii. Chemorefractory relapse.
iii. Relapse after autologous or allogenic transplant.
iv. High risk ALL in CR1: Philadelphia like or any poor risk feature known to be associated with a PFS or DFS ≤40% at 2 years after a conventional transplant.
v. ≥CR2.
vi. MRD+ within 1 month of start of conditioning regimen.
c. Myelodysplastic Syndrome

i. Relapse after allogeneic transplant.
ii. ≥10% blasts within 1 month of start of conditioning regimen.
iii. Very poor risk cytogenetics (> 3 cytogenetic abnormalities).
iv. Any poor risk feature known to be associated with a PFS or DFS ≤40% at 2 years after a conventional transplant.
v. TP53 mutation.
vi. ≥ 40 years old and RAS or JAK-2 mutation.
vii. CMML with HCT-specific CPSS score high or intermediate 2.
viii. Stable disease after 6 cycles of a demethylating agent.
ix. Progressive disease while on a demethylating agent.
2. 18-70 years old.
3. Availability of 2 CBs ≥ 4/6 HLA match when DRB1 is performed at the allele level and A, B at antigen resolution (intermediate resolution) and ≥ 4/8 HLA match when A, B, C and DRB1 are performed at the allele level. An acceptable alternative would be a 3/6 but 5/8 as long as there is no double DRB1 mismatch.

a. Cord to be expanded:

i. CD34+ cell count > 0.5 × 10^5^/kg and TNC > 1.5 × 10^7^/kg (pre-freeze).
ii. Needs to be erythrodepleted by bank prior to cryopreservation.
iii. Should come from a cord bank that is FACT accredited, FDA approved or eligible for NMDP IND.
b. <u>Back up cord</u>: Pre-freeze TNC ≥ 2.0 × 10^7^/kg with CD34^+^ cells ≥ 1.5 × 10^5^/kg or TNC count ≥ 1.5 × 10^7^ TNC/kg with CD34^+^ cells ≥1.7 × 10^5^/kg. If a single cord does not meet these criteria any other acceptable HSC source such as 2 back up cords will be an acceptable alternative (minimum for each of 1.5 × 10^7^ TNC/kg and 1.0 × 10^5^ CD34^+^ cells/kg; an HSC back up source must be available and ready to be collected when conditioning regimen starts (for example: a haploidentical or any other acceptable donor needs to have completed medical clearance prior to start of conditioning regimen).
4. Karnofsky score ≥ 70%.
5. Bilirubin < 2 × upper limit of normal (ULN) unless felt to be related to Gilbert’s disease or hemolysis; AST and ALT ≤ 2.5 × ULN; alkaline phosphatase ≤ 5 × ULN.
6. Estimated or measured creatinine clearance ≥ 60 mL/min/1.73 m^2^.
7. HCT-CI ≤5 for patients <60 years old; HCT-CI ≤3 for patients <60 years old with acute leukemia not in CR/CRi or if 2nd allogeneic transplant; HCT-CI ≤3 for patients who are 60-65 years old; HCT-CI ≤2 for patients 60-65 years old and 2nd transplant or if acute leukemia not in CR/Cri; HCT-CI ≤1 if 66-70 years old; HCT-CI ≤1 and KPS ≥90% if 66-70 years old and 2nd transplant or if acute leukemia not in CR/CRi.
8. Left ventricular ejection fraction ≥ 40% (within 3 months unless patient has received chemotherapy or radiation therapy to the thorax since the last cardiac evaluation).
9. FVC, FEV1 and DLCOc ≥ 50% of predicted (within 3 months unless patient has received chemotherapy or radiation therapy to the thorax since last pulmonary evaluation).
10. Signed written informed consent (parents or legal guardians for minor patients).
11. Female patients of childbearing potential must have a negative serum pregnancy test within 7 days of enrolment and must be willing to use an effective contraceptive method while enrolled in the study.

##### Exclusion criteria

1. Patient never treated with cytotoxic chemotherapy and planned conditioning regimen does not include 12 Gy TBI (exceptions allowed if approved by PI).
2. Allogeneic myeloablative transplant within 6 months.
3. Autologous hematopoietic stem cell transplant within 6 months.
4. Planned use of ATG in conditioning regimen (exceptions allowed if approved by PI in which case ATG must be adjusted for weight/lymphocyte count and given more than 1 week prior to transplant; any patient who receives ATG will have immune recovery studies but will not be counted with rest of patients and will be analyzed separately).
5. Planned use of an HLA matched CB (8/8 allele matched)
6. Uncontrolled infection.
7. HLA antibodies with significant titers directed towards expanded cord blood.
8. Presence of a malignancy other than the one for which the CB transplant is being performed, with an expected survival estimated to be less than 75% at 5 years.
9. Seropositivity for HIV.
10. Hepatitis B or C infection with measurable viral load. Patients with hepatitis B or C infection regardless of viral load require clear documentation of absence of cirrhosis by either fibroscan or biopsy. If fibroscan is the method used, the test must be unequivocally negative.
11. Liver cirrhosis.
12. Active central nervous system involvement.
13. Chloroma > 2 cm.
14. ≥50% blasts in marrow in an evaluable marrow sample (>25% of normal cellularity for age) collected less than one month prior to start of conditioning regimen.
15. Peripheral blasts >1000/mm^3^
16. Pregnancy, breastfeeding or unwillingness to use appropriate contraception.
17. Participation in a trial with an investigational agent within 30 days prior to entry in the study.
18. Patient unable to give informed consent or unable to comply with the treatment protocol including appropriate supportive care, follow-up, and tests.
19. Any abnormal condition or laboratory result that is considered by the PI capable of altering patient’s condition or study outcome.

#### ECT.004

##### Inclusion criteria

1. Age 18 to 70 years old
2. Presence of a high and very high-risk hematologic malignancy defined as:

a. Acute Myeloid Leukemia:

i. Primary induction failure (no CR or CRi after ≥ 2 courses of intensive induction therapy or after ≥ 1 induction containing high dose Ara-C).
ii. Chemorefractory relapse (no CR or CRi after 1 chemointensive treatment).
iii. Relapse after allogeneic or autologous transplant.
iv. High risk AML in CR1 as defined by European Leukemia Net (ELN).
v. ≥ CR2.
vi. Presence of minimal residual disease at the time of transplant.
b. Acute Lymphoid leukemia

i. Primary induction failure (≥ 2 inductions).
ii. High risk ALL in CR1: Ph like ALL128 or any other poor risk feature.
iii. ≥ CR2.
iv. Chemorefractory relapse (at least 1 intensive induction chemotherapy).
v. Relapse after allogeneic or autologous transplant.
vi. Presence of minimal residual disease at the time of transplant.
c. Myelodysplastic syndrome (MDS):

i. Relapse after allogeneic or autologous transplant.
ii. ≥10 % blasts within 30 days of start of conditioning regimen.
iii. Poor and very poor cytogenetics abnormalities.
iv. CMML with HCT-specific CPSS score high or intermediate-2.
v. Stable disease (absence of CR/PR/HI) after 6 cycles of azacitidine (or another demethylating agent).
vi. Progressive disease while on azacitidine (or another demethylating agent).
d. <u>Chronic myelogenous leukemia:</u> Patients who have progressed to blast crisis.
3. Availability of 2 UCBs ≥ 4/6 HLA match when DRB1 is performed at the allele level and A, B at antigen resolution (intermediate resolution) and ≥ 4/8 HLA match when A, B, C and DRB1 are performed at the allele level. An acceptable alternative would be a 3/6 or 5/8 as long as there is no double DRB1 mismatch.

a. Selection of cord to be expanded:

i. Pre-freeze CD34+ cell count ≥ 0.5 × 10^5^/kg and TNC ≥ 1.5 × 10^7^/kg
ii. Needs to be erythrodepleted by bank prior to cryopreservation.
iii. Must comply with local site regulations AND, in the USA, come from a cord bank that is FACT or AABB accredited, FDA approved or eligible for NMDP IND, in Europe come from a cord bank that is FACT, or AABB accredited, or complying with quality standards of the JACIE (Joint Accreditation Committee ISCT-Europe & EBMT) (unless PI approves another bank).
b. Selection of non-expanded CB/back-up cord (recommendation):

i. Pre-freeze CD34+ cells ≥ 1.5 × 10^5^/kg with TNC count ≥ 2.0 × 10^7^/kg or,
ii. Pre-freeze CD34+ cells ≥ 1.7 × 10^5^/kg with TNC count ≥ 1.5 × 10^7^/kg.
iii. If a single cord does not meet these criteria, 2 back up cords will be an acceptable alternative with each having CD34+ cells > 1 × 10^5^/kg with TNC >1.5 × 10^7^/kg.
iv. According to local regulations, must come from a cord bank that is FACT or AABB accredited, or FDA approved or eligible for NMDP IND, or complying with quality standards of the JACIE (unless PI approves another bank).
v. Any other HSC source would be acceptable provided it is available and ready to be collected (e.g., a haploidentical or any other acceptable donor must have completed medical clearance) prior to starting conditioning regimen.
4. Karnofsky ≥70.
5. Left ventricular ejection fraction ≥ 40% (within 3 months unless the patient has received chemotherapy or radiation therapy to the thorax since the last cardiac evaluation) OR fractional shortening >22%.
6. Forced vital capacity (FVC), forced expiratory volume in 1 second (FEV1) and diffusing capacity corrected for hemoglobin (DLCOc) ≥ 50% of predicted.
7. Bilirubin < 2 × upper limit of normal (ULN) unless felt to be related to Gilbert’s disease or hemolysis.
8. AST and ALT ≤ 2.5 × ULN; alkaline phosphatase ≤ 5 × ULN.
9. Adequate renal function defined as creatinine < 2.0 mg/dL. All patients with a creatinine > 1.2 or a history of renal dysfunction must have estimated creatinine clearance > 50 mL/min.
10. Hematopoietic cell transplantation specific comorbidity index (HCT-CI) ≤3 if patients have ≥5% blasts in the bone marrow and HCT-CI ≤5 if 60-70 years old. If a higher comorbidity index is the only non satisfactory eligibility criteria, the Principal Investigator could be consulted to re-evaluate eligibility of this specific patient.

##### Exclusion criteria

1. Allogeneic myeloablative transplant within 6 months.
2. Autologous hematopoietic stem cell transplant within 6 months.
3. Active or recent (prior 6 months) invasive fungal infection without ID consult and approval.
4. The presence of a malignancy other than the one for which the UCB transplant is being performed and the expected survival related to the malignancy is estimated to be less than 75% at 5 years.
5. HIV positivity.
6. Hepatitis B or C infection with measurable viral load.
7. Liver cirrhosis.
8. Pregnancy, breastfeeding or unwillingness to use appropriate contraception.
9. Participation in a trial with an investigational agent within 30 days prior to entry in the study.
10. Patient unable to give informed consent or unable to comply with the treatment protocol including appropriate supportive care, follow-up, and tests.
11. Any abnormal condition or laboratory result that is considered by the principal investigator capable of altering patient condition or study outcome.
12. Active central nervous system involvement.
13. Chloroma > 2 cm.

### Endpoints

#### ECT.002

##### Primary endpoints

1. Evaluate relapse-free survival (RFS) and overall survival (OS) at 1- and 2-years post-transplant. RFS and OS were measured from the time of transplant until progression, death or last follow-up. RFS: an event is defined as relapse or death. OS: an event is defined as death.
2. Confirm a low incidence of transplant-related mortality (TRM) of ≤20% at day 100 and 1 year. TRM was defined as death without prior malignant relapse or recurrence occurring after commencement of the conditioning regimen that could be related to the transplantation procedure. Nonrelated deaths, such as accidental deaths, were excluded.

##### Secondary endpoints

1. Kinetics of hematologic engraftment, including time to neutrophil and platelet engraftment, and incidence of primary and late graft failure;
2. incidence of acute and chronic GVHD by NIH criteria at 2 years post-transplant;
3. incidence of grade 3 or higher infectious complications;
4. incidence of hospitalization events;
5. incidence of pre-engraftment/engraftment syndrome requiring therapy;

##### Exploratory endpoints

1. Immune reconstitution, including T-, B-, NK-. dendritic cell reconstitution;
2. identification of predictors of early-onset alloimmune reactions and their influence on TRM and relapse;
3. influence of HLA and killer-cell immunoglobulin-like receptor ligand mismatch on transplantation outcomes;
4. relationship between mycophenolate levels and GVHD, TRM, and relapse;
5. sequencing of leukemia/myelodysplasia that relapsed to identify markers suggesting escape from the immune system;
6. graft composition;
7. evaluation of mucositis by measurement of bacterial translocation using gene sequencing in blood, blood citrulline levels, and number of days of parenteral nutrition;
8. cost evaluation;
9. relapse and survival between 3 and 5 years post-transplant;
10. bank of biological samples

#### ECT.004

##### Primary endpoints

1. Evaluate relapse-free survival (RFS) and overall survival (OS) at 1- and 2-years post-transplant. RFS and OS were measured from the time of transplant until progression, death or last follow-up. RFS: an event is defined as relapse/progression or death. OS: an event is defined as death.
2. Examine the safety, including assessment of the rate of graft failure, and feasibility of infusing a single ECT-001-expanded cord blood unit. Safety was assessed primarily by the tabulation of adverse events and review of laboratory evaluations. Adverse events were graded according to the modified National Cancer Institute Common Terminology Criteria for Adverse Events (CTCAE), version 5.0. All grade 3–5 toxicities were compiled up to discharge from the transplant center and thereafter at the 6-, 12-, and 24-month time points.

##### Secondary endpoints

1. Kinetics of hematologic engraftment (time to neutrophil and platelet engraftment);
2. estimate the incidence of transplant related mortality at day 100 and 1-year post-transplant;
3. incidence of acute and chronic GVHD by NIH criteria at 2 years post-transplant;
4. evaluate GRFS and CRFS at 1- and 2-years post-transplantation;
5. incidence of grade 3 or higher infectious complications;
6. incidence of pre-engraftment/engraftment syndrome requiring therapy

##### Exploratory endpoints

1. Immune reconstitution;
2. Hospitalization events, including duration of transplant admission and number of days in hospital in 1st 100 days, last day of fever (≥38°C) prior to engraftment;
3. Dendritic and mast cell analysis in patient;
4. Detailed cellular and molecular analysis of graft at single cell level;
5. Sequencing of leukemias/myelodysplasias that relapse to identify markers suggesting an escape from the immune system
6. QoL evaluation (EU sites).

### Manufacturing details of UM171 expanded cord blood

The selected cryopreserved CB unit was initially thawed and subjected to CD34+ enrichment using the CD34+ CliniMACS selection system (Miltenyi). After CD34+ selection, the CD34-lymphocyte containing fraction, called Drug Product 2 (DP2), was cryopreserved to be infused at transplantation. The CD34+ fraction was inoculated in gas permeable cell culture bags and subjected to a 7-day culture in a fed-batch system. Briefly, a small volume of serum- and animal component-free medium supplemented with GMP-grade cytokines (SCF, IL6, TPO, FLT3) and UM171 (full proprietary name: UM0128171), was added daily to the culture bag. Cells were gently mixed on an orbital shaker and maintained at a 37°C and 5% CO2 in a cell culture incubator. The small culture volume (median ≈700 mL) and the robustness of the optimized culture process decreased the risk of contamination. After 7 days, the cell suspension was washed, formulated and tested. During washing, all UM171 is removed. The expanded product, called Drug Product 1 (DP1) was infused fresh in 2 patients and cryopreserved for later in infusion in 58 patients. UM171 grafts were manufactured at the Centre d’Excellence en Thérapie Cellulaire (CETC) at Maisonneuve Rosemont Hospital (Montréal).

### Expanded cord blood release criteria

Prior to CB manipulation, the CB unit was required to have total nucleated cell (TNC) viability ≥40% after thaw. The manufacturing process always started with a single CB unit obtained from a public CB bank and resulted in two drug products: DP1 (the CD34+ expanded component) and DP2 (the CD34-fraction). The CD34-negative fraction (DP2) had to contain a minimal dose of 1.0 × 10^6^ CD3+ cells/kg after CD34+ selection and before cryopreservation. The expanded CD34+ fraction (DP1) had to contain a minimum of 5.0 × 10⁵ CD34+ cells/kg, have a CD34+ cell viability of 70% or more, and have negative microbiology testing (i.e., culture, endotoxin, and PCR mycoplasma).

### Post transplant monitoring

Safety was assessed by tabulation of grade 3-5 adverse events and the review of laboratory evaluations compiled on a continuous basis during hospitalisation and regularly thereafter at 3, 6, 12, and 24 months (until 36 months in study ECT-002). As prespecified per protocol, all grade 3-5 events were collected and evaluated according to the NCI Common Terminology Criteria for Adverse Events (CTCAE) version 4.0 (grade 1-2 events were not recorded). However, ECT-004 did not capture expected grade 3-5 AEs associated with HCT.

Patients had routine post-transplant monitoring as per local institution standards (e.g., blood counts/chemistry, medical visits, viral monitoring; CD4+ counts were performed only in ECT-002). Bone marrow sampling was performed at 3 and 12 months (as well at 1 month on ECT-004). Chimerism studies were done using short variable tandem repeats by PCR assay for specific cell populations (CD3+ for T cells and CD33+ for myeloid cells) on days 14, 28 and at 3, 6 and 12 months. Patients were followed until 2 years post transplant on study ECT-004 and 3 years on study ECT-002.

## Supplementary Tables

**Supplementary Table 1.** Summary of Non-Relapse Mortality (NRM) Events and Causes.

| **NRM#** | **NRM**<br>**occurrence**<br>**(months)** | **Patient**<br>**Age-range**<br>**(years)** | **Primary cause of death** | **Contributing cause(s)** |
| --- | --- | --- | --- | --- |
| 1 | 0.6 | 45-49 | Citrobacter Sepsis | Enterococcal Infection |
| 2 | 0.9 | 60-64 | Idiopathic Pneumonia Syndrome | - |
| 3 | 1.4 | 45-49 | Respiratory Failure | Graft failure |
| 4 | 4.6 | 40-44 | Metapneumovirus Infection | - |
| 5 | 5.3 | 50-54 | Sepsis | COVID-19 |
| 6 | 6.2 | 55-59 | Pneumonia (aspergillosis) | - |
| 7 | 6.7 | 55-59 | Multiple Organ Dysfunction Syndrome | - |
| 8 | 7.7 | 55-59 | Pulmonary Hypertension | - |
| 9 | 9.1 | 60-64 | Organising Pneumonia | - |
| 10 | 13.2 | 55-59 | Pneumonia | Refractory acute GVHD |
| 11 | 13.6 | 40-44 | Septic Shock | - |
| 12 | 21.9 | 50-54 | Respiratory Failure | - |

**Supplementary Table 2.** Summary of grade ≥ 3 Adverse Events.

| Preferred Term | Grade 3 | Grade 4 | Grade 5 | All |
| --- | --- | --- | --- | --- |
| **Number of participants with any AEs** | **18 (30.0%)** | **29 (48.3%)** | **12 (20.0%)** | **59 (98.3%)** |
| Blood And Lymphatic System Disorders - ECT-002 only (n=30)* |  |  |  |  |
| Lymphopenia | 1 (3.3%) | 29 (96.7%) | - | 30 (100.0%) |
| Neutropenia | - | 30 (100.0%) | - | 30 (100.0%) |
| Thrombocytopenia | - | 30 (100.0%) | - | 30 (100.0%) |
| Anaemia | 27 (90.0%) | 2 (6.7%) | - | 29 (96.7%) |
| Leukopenia | - | 29 (96.7%) | - | 29 (96.7%) |
| Febrile Neutropenia | 13 (43.3%) | - | - | 13 (43.3%) |
| All other SOC -ECT-002 and ECT-004 (n=60) |  |  |  |  |
| Decreased appetite / Malnutrition | 26 (43.3%) | - | - | 26 (43.3%) |
| Hyperglycaemia | 21 (35.0%) | - | - | 21 (35.0%) |
| Nausea / Vomiting | 19 (31.7%) | - | - | 19 (31.7%) |
| Hypertension | 18 (30.0%) | - | - | 18 (30.0%) |
| Pneumonia | 14 (23.3%) | - | 2 (3.3%) | 16 (26.7%) |
| Sepsis / Bacteraemia / Viraemia | 14 (23.3%) | - | 2 (3.3%) | 16 (26.7%) |
| Diarrhoea | 14 (23.3%) | - | - | 14 (23.3%) |
| Mucosal inflammation | 12 (20.0%) | - | - | 12 (20.0%) |
| Acute graft versus host disease# | 9 (15.0%) | 1 (1.7%) | 1 (1.7%) | 11 (18.3%) |
| COVID-19 | 7 (11.7%) | - | 1 (1.7%) | 8 (13.3%) |
| Cytomegalovirus reactivation | 8 (13.3%) | - | - | 8 (13.3%) |
| Hypogammaglobulinaemia | 8 (13.3%) | - | - | 8 (13.3%) |
| Acute kidney injury / Renal failure | 6 (10.0%) | 1 (1.7%) | - | 7 (11.7%) |
| Alanine / aspartate aminotransferase increased | 6 (10.0%) | 1 (1.7%) | - | 7 (11.7%) |
| Engraftment syndrome | 7 (11.7%) | - | - | 7 (11.7%) |
| Epstein-Barr Virus Infection | 7 (11.7%) | - | - | 7 (11.7%) |
| Headache | 7 (11.7%) | - | - | 7 (11.7%) |
| Myopathy / Muscular weakness | 7 (11.7%) | - | - | 7 (11.7%) |
| Rash maculo-papular | 7 (11.7%) | - | - | 7 (11.7%) |
| Hypervolaemia | 6 (10.0%) | - | - | 6 (10.0%) |
| Malaise / Fatigue | 6 (10.0%) | - | - | 6 (10.0%) |
| Abdominal pain / Abdominal pain upper | 5 (8.3%) | - | - | 5 (8.3%) |
| Device related infection | 5 (8.3%) | - | - | 5 (8.3%) |
| Pyrexia | 5 (8.3%) | - | - | 5 (8.3%) |
| Syncope | 5 (8.3%) | - | - | 5 (8.3%) |
| Blood creatinine increased | 4 (6.7%) | - | - | 4 (6.7%) |
| Delirium | 4 (6.7%) | - | - | 4 (6.7%) |
| Depression /anxiety / suicide attempt | 3 (5.0%) | 1 (1.7%) | - | 4 (6.7%) |
| Electrocardiogram qt prolonged | 4 (6.7%) | - | - | 4 (6.7%) |
| Human herpesvirus 6 infection | 3 (5.0%) | 1 (1.7%) | - | 4 (6.7%) |
| Hypotension | 4 (6.7%) | - | - | 4 (6.7%) |
| Respiratory tract infection / Upper respiratory tract infection | 4 (6.7%) | - | - | 4 (6.7%) |
| Adenovirus infection | 3 (5.0%) | - | - | 3 (5.0%) |
| Adenovirus reactivation | 3 (5.0%) | - | - | 3 (5.0%) |
| Bone pain | 3 (5.0%) | - | - | 3 (5.0%) |
| Hypoalbuminaemia | 3 (5.0%) | - | - | 3 (5.0%) |
| Hypokalaemia | 3 (5.0%) | - | - | 3 (5.0%) |
| Hypomagnesaemia | 3 (5.0%) | - | - | 3 (5.0%) |
| Idiopathic pneumonia syndrome | 2 (3.3%) | - | 1 (1.7%) | 3 (5.0%) |
| Iron overload | 3 (5.0%) | - | - | 3 (5.0%) |
| Organising pneumonia | 1 (1.7%) | 1 (1.7%) | 1 (1.7%) | 3 (5.0%) |
| Pain in extremity | 3 (5.0%) | - | - | 3 (5.0%) |
| Peripheral neuropathy | 3 (5.0%) | - | - | 3 (5.0%) |
| Anaphylactic reaction | 2 (3.3%) | - | - | 2 (3.3%) |
| Back pain | 2 (3.3%) | - | - | 2 (3.3%) |
| Bronchopulmonary aspergillosis | 2 (3.3%) | - | - | 2 (3.3%) |
| Cardiac failure | 2 (3.3%) | - | - | 2 (3.3%) |
| Cataract | 2 (3.3%) | - | - | 2 (3.3%) |
| Cellulitis | 2 (3.3%) | - | - | 2 (3.3%) |
| Clostridium difficile colitis | 2 (3.3%) | - | - | 2 (3.3%) |
| Enterococcal infection | 1 (1.7%) | - | 1 (1.7%) | 2 (3.3%) |
| Epistaxis | 2 (3.3%) | - | - | 2 (3.3%) |
| Haemolytic uraemic syndrome | 2 (3.3%) | - | - | 2 (3.3%) |
| Herpes zoster | 2 (3.3%) | - | - | 2 (3.3%) |
| Hypoglycaemia | 2 (3.3%) | - | - | 2 (3.3%) |
| Intestinal obstruction / ileus | 2 (3.3%) | - | - | 2 (3.3%) |
| Malabsorption | 2 (3.3%) | - | - | 2 (3.3%) |
| Microangiopathy | 2 (3.3%) | - | - | 2 (3.3%) |
| Multiple organ dysfunction syndrome | - | 1 (1.7%) | 1 (1.7%) | 2 (3.3%) |
| Pericardial effusion | 2 (3.3%) | - | - | 2 (3.3%) |
| Respiratory failure | - | - | 2 (3.3%) | 2 (3.3%) |
| Staphylococcal bacteraemia | 2 (3.3%) | - | - | 2 (3.3%) |
| Stomatitis | 2 (3.3%) | - | - | 2 (3.3%) |
| Transplant failure | - | 2 (3.3%) | - | 2 (3.3%) |
| Urinary tract infection | 2 (3.3%) | - | - | 2 (3.3%) |
| Venoocclusive liver disease | 2 (3.3%) | - | - | 2 (3.3%) |
| Acute myocardial infarction | 1 (1.7%) | - | - | 1 (1.7%) |
| Anal fissure | 1 (1.7%) | - | - | 1 (1.7%) |
| Angina pectoris | 1 (1.7%) | - | - | 1 (1.7%) |
| Appendicitis | 1 (1.7%) | - | - | 1 (1.7%) |
| Aspiration | 1 (1.7%) | - | - | 1 (1.7%) |
| Asthma | 1 (1.7%) | - | - | 1 (1.7%) |
| Atrial flutter | 1 (1.7%) | - | - | 1 (1.7%) |
| Bk virus infection | 1 (1.7%) | - | - | 1 (1.7%) |
| Blood alkaline phosphatase increased | 1 (1.7%) | - | - | 1 (1.7%) |
| Blood lactate dehydrogenase increased | 1 (1.7%) | - | - | 1 (1.7%) |
| Campylobacter colitis | 1 (1.7%) | - | - | 1 (1.7%) |
| Carbon monoxide diffusing capacity decreased | 1 (1.7%) | - | - | 1 (1.7%) |
| Citrobacter sepsis | - | - | 1 (1.7%) | 1 (1.7%) |
| Colitis | 1 (1.7%) | - | - | 1 (1.7%) |
| Complication associated with device | 1 (1.7%) | - | - | 1 (1.7%) |
| Cystitis haemorrhagic | 1 (1.7%) | - | - | 1 (1.7%) |
| Cytogenetic abnormality | 1 (1.7%) | - | - | 1 (1.7%) |
| Dermatitis acneiform | 1 (1.7%) | - | - | 1 (1.7%) |
| Diabetes mellitus | 1 (1.7%) | - | - | 1 (1.7%) |
| Dysphagia | 1 (1.7%) | - | - | 1 (1.7%) |
| Ejection fraction decreased | 1 (1.7%) | - | - | 1 (1.7%) |
| Encephalopathy | 1 (1.7%) | - | - | 1 (1.7%) |
| Enterocolitis | 1 (1.7%) | - | - | 1 (1.7%) |
| Extremity necrosis | 1 (1.7%) | - | - | 1 (1.7%) |
| Flank pain | 1 (1.7%) | - | - | 1 (1.7%) |
| Food poisoning | 1 (1.7%) | - | - | 1 (1.7%) |
| Gastritis | 1 (1.7%) | - | - | 1 (1.7%) |
| Gastrointestinal candidiasis | 1 (1.7%) | - | - | 1 (1.7%) |
| Generalised oedema | 1 (1.7%) | - | - | 1 (1.7%) |
| Haematuria | 1 (1.7%) | - | - | 1 (1.7%) |
| Hepatitis | 1 (1.7%) | - | - | 1 (1.7%) |
| Hiccups | 1 (1.7%) | - | - | 1 (1.7%) |
| Hyperbilirubinaemia | 1 (1.7%) | - | - | 1 (1.7%) |
| Hyperkalaemia | 1 (1.7%) | - | - | 1 (1.7%) |
| Hypermagnesaemia | 1 (1.7%) | - | - | 1 (1.7%) |
| Hypovolaemia | 1 (1.7%) | - | - | 1 (1.7%) |
| Hypoxia | 1 (1.7%) | - | - | 1 (1.7%) |
| Immunoglobulins decreased | 1 (1.7%) | - | - | 1 (1.7%) |
| Infective myositis | 1 (1.7%) | - | - | 1 (1.7%) |
| Jejunal perforation | - | 1 (1.7%) | - | 1 (1.7%) |
| Klebsiella sepsis | 1 (1.7%) | - | - | 1 (1.7%) |
| Lipodystrophy acquired | 1 (1.7%) | - | - | 1 (1.7%) |
| Metabolic encephalopathy | - | 1 (1.7%) | - | 1 (1.7%) |
| Metapneumovirus infection | 0 (0.0%) | - | 1 (1.7%) | 1 (1.7%) |
| Mouth haemorrhage | 0 (0.0%) | 1 (1.7%) | - | 1 (1.7%) |
| Muscle spasms | 1 (1.7%) | - | - | 1 (1.7%) |
| Myalgia | 1 (1.7%) | - | - | 1 (1.7%) |
| Myocardial infarction | 1 (1.7%) | - | - | 1 (1.7%) |
| Nephrolithiasis | 1 (1.7%) | - | - | 1 (1.7%) |
| Non-cardiac chest pain | 1 (1.7%) | - | - | 1 (1.7%) |
| Oedema peripheral | 1 (1.7%) | - | - | 1 (1.7%) |
| Oesophageal candidiasis | 1 (1.7%) | - | - | 1 (1.7%) |
| Oesophagitis | 1 (1.7%) | - | - | 1 (1.7%) |
| Orthostatic hypotension | 1 (1.7%) | - | - | 1 (1.7%) |
| Pelvic pain | 1 (1.7%) | - | - | 1 (1.7%) |
| Pericarditis | 1 (1.7%) | - | - | 1 (1.7%) |
| Pleuritic pain | 1 (1.7%) | - | - | 1 (1.7%) |
| Pneumatosis intestinalis | 1 (1.7%) | - | - | 1 (1.7%) |
| Pneumocystis jirovecii pneumonia | 1 (1.7%) | - | - | 1 (1.7%) |
| Pruritus | 1 (1.7%) | - | - | 1 (1.7%) |
| Pulmonary embolism | 1 (1.7%) | - | - | 1 (1.7%) |
| Pulmonary hypertension | - | - | 1 (1.7%) | 1 (1.7%) |
| Roseolovirus test positive | 1 (1.7%) | - | - | 1 (1.7%) |
| Scan abnormal | 1 (1.7%) | - | - | 1 (1.7%) |
| Serum ferritin increased | 1 (1.7%) | - | - | 1 (1.7%) |
| Sinusitis | 1 (1.7%) | - | - | 1 (1.7%) |
| Soft tissue necrosis | 1 (1.7%) | - | - | 1 (1.7%) |
| Streptococcal sepsis | 1 (1.7%) | - | - | 1 (1.7%) |
| Thrombophlebitis septic | 1 (1.7%) | - | - | 1 (1.7%) |
| Thrombotic microangiopathy | 1 (1.7%) | - | - | 1 (1.7%) |
| Transfusion reaction | 1 (1.7%) | - | - | 1 (1.7%) |
| Vascular access complication | 1 (1.7%) | - | - | 1 (1.7%) |
| Vulvitis | 1 (1.7%) | - | - | 1 (1.7%) |

AEs were classified into SOC and preferred term using MedDRA Version 23.1 (or later).
A participant with multiple events coding to the same preferred term within a primary SOC was counted only once for the preferred term within that primary SOC.
\*: include the occurrence of intended events reported in the investigator-initiated Study 002 protocol.

## Supplementary Figures

**Supplementary Figure 1.**
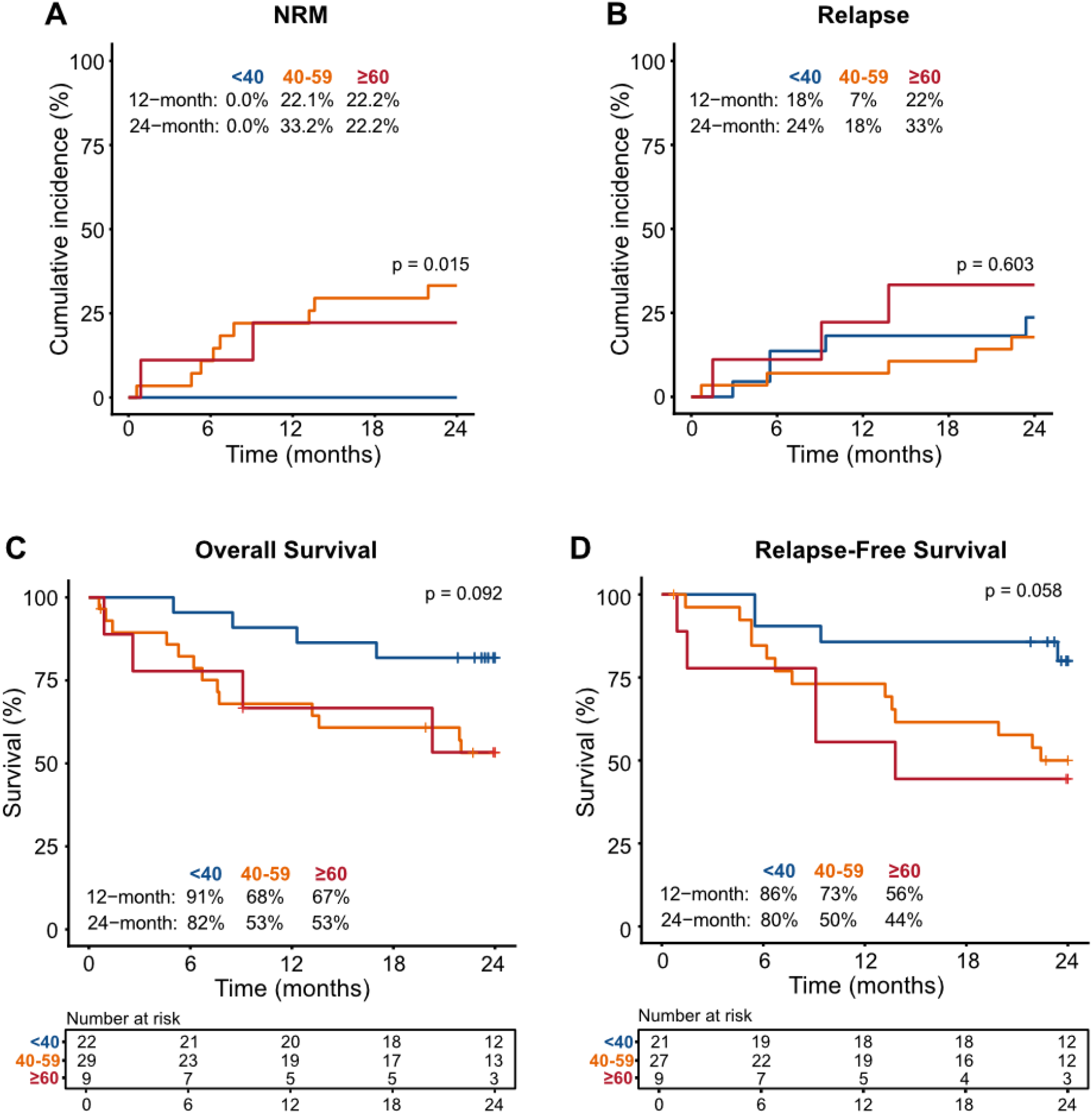
Non-relapse mortality, relapse and survival outcomes following UM171-expanded cord blood transplantation stratified by age (< 40, 40-59, and ≥ 60 years). (A-B) Cumulative incidence of NRM (A) and relapse (B). (C-D) Kaplan-Meier estimates of OS (C) and RFS (D). Tick marks indicate censored observations. Numbers at risk are shown below each panel. Gray’s test was used for comparisons of cumulative incidence functions, and the log-rank test for Kaplan-Meier curves.

**Supplementary Figure 2.**
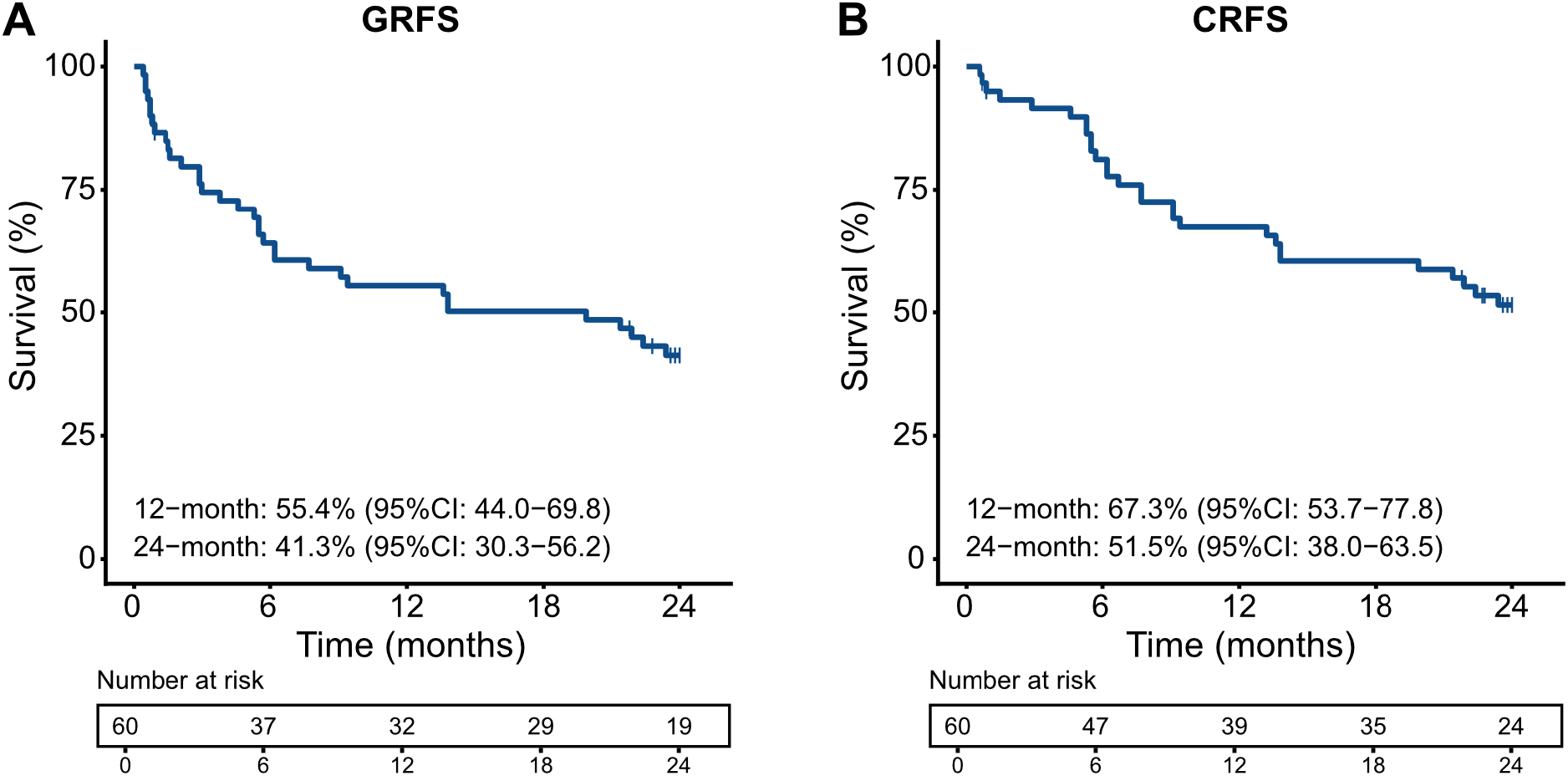
Composite outcomes following UM171-expanded cord blood transplantation. (A) GVHD-free, relapse-free survival (GRFS). (B) Chronic GVHD–free, relapse-free survival (CRFS). Kaplan-Meier estimates are shown, with tick marks indicating censored observations. Twelve- and 24-month estimates with 95% confidence intervals are provided. Numbers at risk are shown below each panel.

**Supplementary Figure 3.**
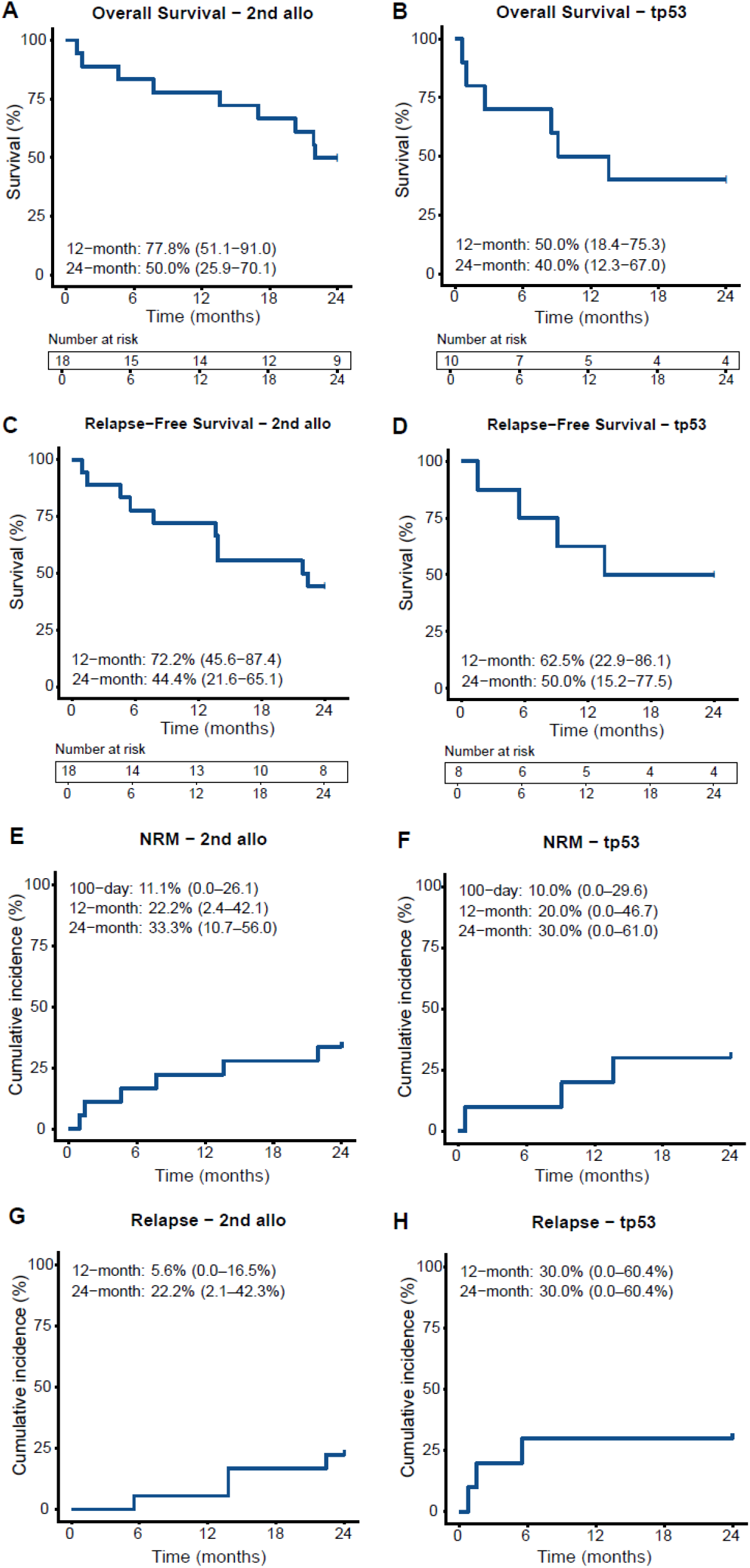
Outcomes in high-risk subgroups: second allogeneic transplantation and TP53-mutated disease. (A) Overall survival (OS) in patients undergoing a second allogeneic transplantation. (B) OS in patients with TP53-mutated disease. (C) Relapse-free survival (RFS) in patients undergoing a second allogeneic transplantation. (D) RFS in patients with TP53-mutated disease. Kaplan-Meier estimates are shown, with tick marks indicating censored observations and numbers at risk displayed below each panel. Twelve- and 24-month estimates are provided. (E) Cumulative incidence of non-relapse mortality (NRM) in patients undergoing a second allogeneic transplantation. (F) NRM in patients with TP53-mutated disease. (G) Cumulative incidence of relapse in patients undergoing a second allogeneic transplantation. (H) Relapse in patients with TP53-mutated disease. Cumulative incidence functions are shown.

